# Transmission of aerosols through pristine and reprocessed N95 respirators

**DOI:** 10.1101/2020.05.14.20094821

**Authors:** Paul Z. Chen, Aldrich Ngan, Niclas Manson, Jason T. Maynes, Gregory H. Borschel, Ori D. Rotstein, Frank X. Gu

## Abstract

During the Covid-19 pandemic, pristine and reprocessed N95 respirators are crucial equipment towards limiting nosocomial infections. The NIOSH test certifying the N95 rating, however, poorly simulates aerosols in healthcare settings, limiting our understanding of the exposure risk for healthcare workers wearing these masks, especially reprocessed ones. We used experimental conditions that simulated the sizes, densities and airflow properties of infectious aerosols in healthcare settings. We analyzed the penetration and leakage of aerosols through pristine and reprocessed N95 respirators. Seven reprocessing methods were investigated. Our findings suggest that pristine and properly reprocessed N95 respirators effectively limit exposure to infectious aerosols, but that care must be taken to avoid the elucidated degradation mechanisms and limit noncompliant wear.

---

During the Covid-19 pandemic, disposable N95 filtering facepiece respirators (N95 FFRs) are crucial equipment towards limiting nosocomial infections.^1^ To address critical shortages, reprocessing is being implemented to facilitate their limited reuse.^2^ The N95 rating suggests that up to 5% of airborne particles may transmit through an N95 FFR. The NIOSH tests certifying this rating, however, poorly simulate the transmission of aerosols in healthcare settings,^3^ limiting our understanding of the exposure risk for healthcare workers performing aerosol-generating medical procedures and of the implications of reprocessing.

We analyzed the penetration (transmission through the filter media) and leakage (transmission around imperfections in facial seal) of aerosols into pristine and reprocessed N95 FFRs. We examined three prevalent healthcare models (3M 1860S, 3M 8210 and 3M 9210) and reprocessed them (1, 3, 5 or 10 cycles) using seven methods under consideration for implementation in hospitals: autoclave, 70% ethanol vapor (vEtOH), forced-air dry heat (100 °C), humid heat (75% relative humidity, 75 °C), hydrogen peroxide gas plasma (HPGP, STERRAD® 100S), hydrogen peroxide vapor (HPV, STERIS V-PRO®) and ultraviolet germicidal irradiation (UVGI). Leakage was assessed via fit testing. Penetration was evaluated using a polydisperse challenge aerosol (0.1 to 1 μm; material density, 1.05 g/cm^3^) and conditions that simulated the sizes, densities and airflow properties of infectious aerosols in healthcare settings (see the Supplementary Appendix for the Experimental design and Methods sections).^4^

For both pristine (Fig. 1A) and reprocessed (Fig. S1 to S7) N95 FFRs, penetration rapidly decreased as aerosol size increased according to a power relationship. Power regression verified this trend, with a cumulative *R*^2^ of 0.94 ± 0.041 for all penetration experiments in this study (*N* = 72). For pristine N95 FFRs, the expected aerosol penetration was between 0.09% and 0.19% at 0.1 μm, 0.02% and 0.03% at 0.3 μm and at the detection limit, 0.01%, above 0.5 μm (Fig. 1A).

**Figure 1.**
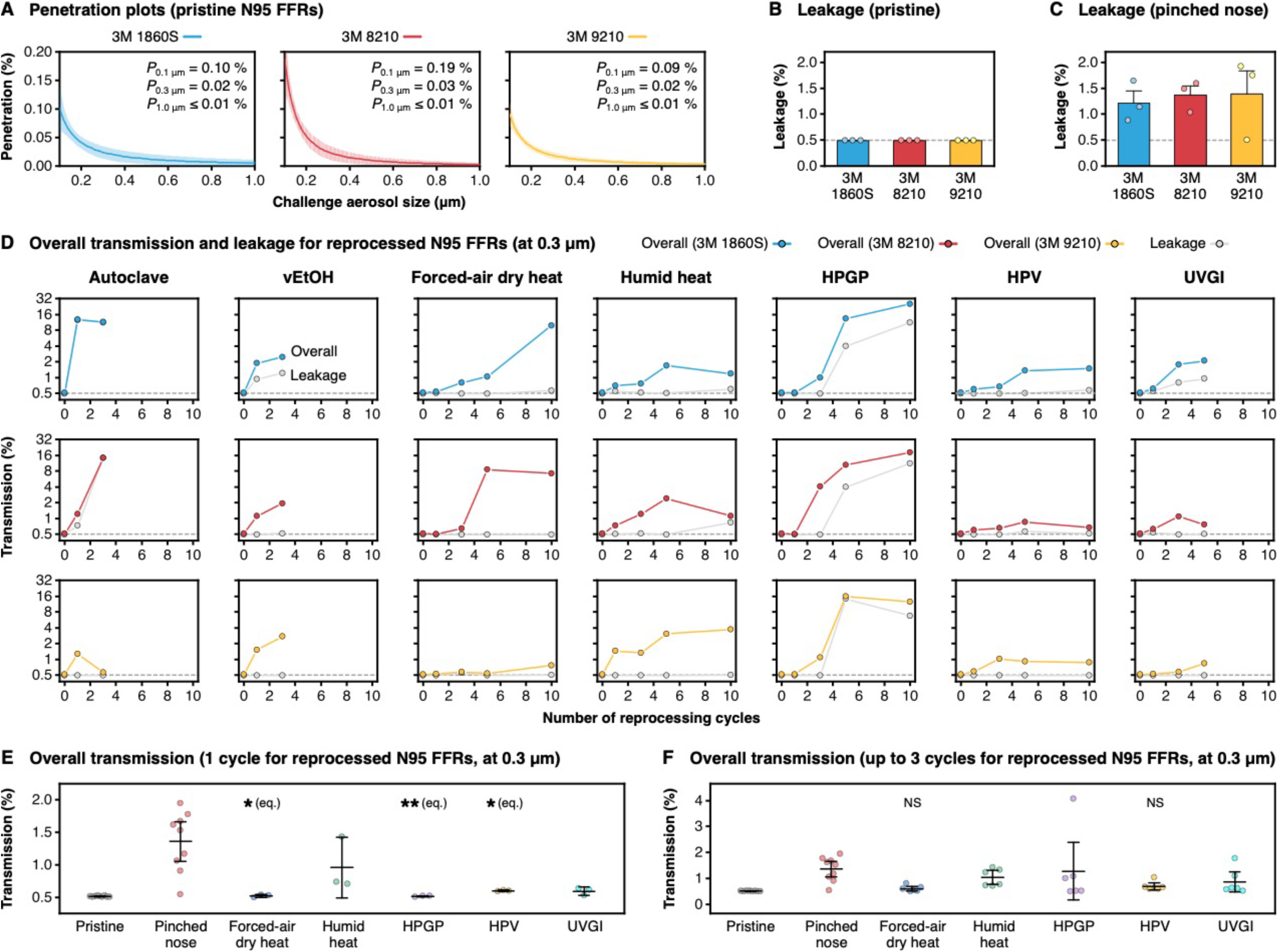
Penetration, leakage and overall transmission of aerosols into pristine and reprocessed N95 FFRs. (**A**) Aerosol penetration was measured using three models of pristine N95 FFRs. A polydisperse challenge aerosol (0.1 to 1.0 μm, material density of 1.05 g/cm^3^) was introduced at 1.0 scfm while experimental conditions were maintained at 20.9 ± 0.52 °C and 48.5 ± 3.70% RH. Power regression was performed on discrete measurements. Plots show the size-dependent expectation curves and the 95% confidence intervals of individual samples. The inset values represent the expected penetration at 0.1, 0.3 and 1.0 μm. The detection limit was 0.01% across aerosol sizes. (**B** and **C**) Aerosol leakage was quantified into three models of pristine N95 FFRs worn properly (B) or with a pinched nose clip (C). Graphs show size-independent means and their standard errors. The dashed lines indicate the detection limit, which was 0.49%. (**D**) The three models of N95 FFRs were reprocessed for 1, 3, 5 or 10 cycles and characterized for penetration and leakage while worn properly. We only implemented autoclave and vEtOH up to 3 cycles; and UVGI up to 5 cycles. Plots show the overall aerosol transmission, the sum of the expected penetration and leakage, at 0.3 μm and leakage over cycle numbers. The dashed lines indicate the detection limit for overall transmission, which was 0.50%. (**E** and **F**) Pristine N95 FFRs were compared with ones reprocessed up to 1 cycle (E) or 3 cycles (F). Plots show the individual data points, means and 95% confidence intervals for overall aerosol transmission at 0.3 μm. Data across N95 models were grouped together up to each cycle number. Autoclave and vEtOH were excluded from these plots and reported in the Supplementary Appendix. Equivalence testing compared reprocessed N95 FFRs with pristine ones. *\*P* < 0.01, *\*\*P* < 0.001; NS, non-significant (*P* > 0.05).

When properly fitted, we measured aerosol leakage at the detection limit, 0.49%, for the three N95 models (Fig. 1B). As such, the estimated overall transmission (sum of penetration and leakage) was ≤0.68%, with the most penetrating particle size at 0.1 μm. Improper wear due to a pinched nose clip, a common issue, significantly enhanced leakage (Fig. 1C).

Reprocessing methods varied in their effects on aerosol transmission (Fig. 1D). At 0.3 μm, HPV kept overall transmission below 1.5% up to 10 cycles, while force-air dry heat and humid heat did so up to 3 cycles. HPGP and UVGI did for 1 cycle but increased transmission by the third cycle; samples reprocessed twice were not included in this study. These five methods kept leakage below 0.6% for the identified cycles. Autoclave physically deformed the pleated models (3M 1860S and 3M 8210), inducing leakage; the molded model (3M 9210) was unaffected. UVGI induced slight dose-dependent photochemical damage (Fig. S8). HPGP caused leakage around the nose by 5 cycles: reactive oxygen species generated during the plasma phase progressively embrittled and degraded polyurethane nose foams across N95 models (Fig. S9 and S10). For mechanistic insight into how reprocessing increased penetration, we measured the pressure differential, which indicates structural changes, across each FFR. N95 filter media collect aerosols based on their static charge or structure. Pressure differentials stayed consistent (Table S1), implying the seven methods increased penetration mainly by degrading filter charge.

Equivalence testing demonstrated that N95 FFRs reprocessed once using forced-air dry heat, HPGP or HPV were statistically equivalent to pristine ones in terms of aerosol transmission (Fig. 1E and S11, *P* < 0.01 or *P* < 0.001), subverting the conventional expectation that the very act of reprocessing increases transmission. No N95 FFRs showed equivalency up to 3 cycles (Fig. 1F and S12, *P* > 0.05).

Our findings help better understand aerosol exposure for healthcare workers wearing N95 FFRs. Since the size of SARS-CoV-2 and influenza virions is approximately 0.1 μm,^5^ infectious aerosols containing them are larger than 0.1 μm. Our results suggest that <0.68% of these virus-containing aerosols transmit into a pristine N95 FFR. Our data indicates HPV, forced-air dry heat, humid heat, HPGP and UVGI maintain <1.5% transmission at 0.3 μm, and in some cases preserve pristine performance, within the identified cycle numbers. The established power relationship demonstrates penetration decreases considerably as aerosol size increases. In comparison, improper wear induces significant leakage, highlighting the importance of compliant wear. These findings suggest pristine and properly reprocessed N95 FFRs effectively protect against infectious aerosols, but that care must be taken during use and reprocessing to mitigate degradation of filter charge, avoid deterioration of straps and nose foams, preserve mask shape especially for molded models and limit noncompliant wear.

## Data Availability

The authors confirm that the data supporting the findings of this study are available within the article and its supplementary materials.

## Supplementary Appendix

### Experimental design

We designed this study to better understand how aerosols relevant to nosocomial infections transmit through N95 FFRs. The N95 rating means that the FFR filter is not resistant to oil and that a minimum of 95% of airborne particles are filtered while fitted properly. We chose not to use the NIOSH certification tests (42 CFR Part 84, TEB-APR-STP-0059 protocol) for our study.^3,6^ This protocol characterizes filtration efficiency by using relatively monodisperse 75-nm NaCl particles (material density, 2.16 g/cm^3^), using specific humidity conditions (preconditioning at 85% relative humidity and 38 °C for 24 h) and loading particulate matter up to the mass threshold (200 mg) under increased flow rates (85 L/min).^6^ In healthcare environments, it is not expected for N95 FFRs to uptake particulates up to the loading threshold.^7^ Moreover, as explained below, the particulates and conditions used in this testing protocol are dissimilar to the aerosols and conditions of interest for this study.

The airflow and transmission characteristics of aerosols depend on the physicochemical properties of the aerosol and the properties of the surrounding gas. Description of the motion of spherical aerosols can be formalized by the Maxey and Riley differential force balance, the relative Reynolds number (Re), Stokes’ law and a statistical treatment of Brownian motion.^8-11^ The differential force equation can be written in the *x*-direction in Cartesian coordinates as

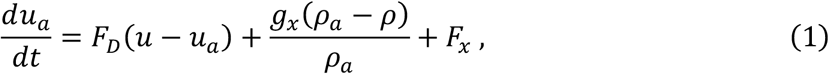

where *u_a_* is the aerosol particle velocity, *t* is time, ***F****_D_* (***u*** − *u_a_*) is the drag force per unit particle mass, *ρ_a_* is the aerosol particle material density, *ρ* is the fluid (in our case, gas) material density and *F_x_* accounts for additional forces acting on the system. The relative Reynolds number is defined as

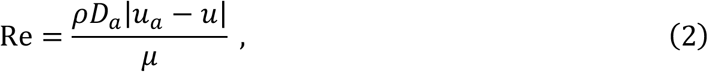

where *D_a_* is the aerosol particle diameter, *u* is the fluid velocity and *μ* is the dynamic viscosity of the fluid. For submicron aerosol particles, Stokes’ law describes

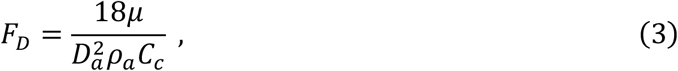

where the Cunningham correction factor is defined as

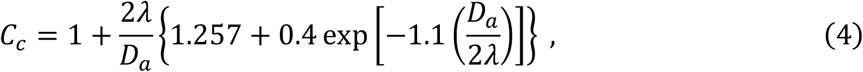

where *λ* is the molecular mean-free path of the aerosol particle. When including the forces required to accelerate the fluid surrounding the particle and due to a pressure gradient in the fluid, the additional force term in eq. (1) can written as

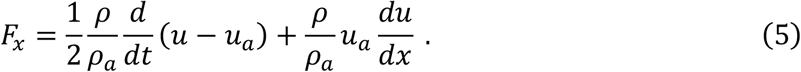

The effects of Brownian motion, which are important for smaller aerosols, can be included as well. The amplitudes of the Brownian forces components are described by

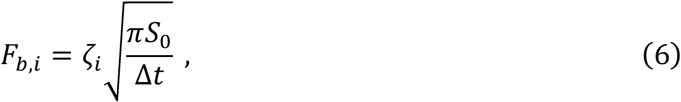

where ζ*_i_* are zero-mean, unit-variance-independent Gaussian random numbers at time step *i*. The components of the Brownian forces can be modeled as a Gaussian white noise process with spectra intensity 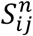 defined as

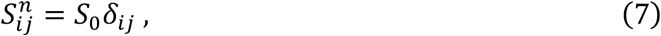

where *δ_ij_*, is the Kronecker delta and

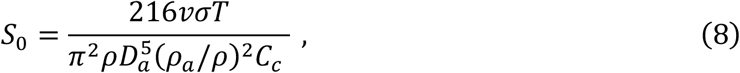

where *T* is the absolute temperature and *v* is the kinematic viscosity.

Taken together, eqs. (1) to (8) show that airflow and transmission characteristics of aerosols depend on the size, material density, surface charge and morphology of the aerosol as well as the composition, flow and temperature of the surrounding gas.

While, at the time of writing,^12^ Covid-19 is believed to be communicated through the droplet and contact modes of transmission,^13,14^ aerosol-generating medical procedures (AGMPs) discharge aerosols (conventionally, <5 μm), potentially leading to nosocomial infection.^15^ Other infectious diseases, such as influenza, can induce respiratory infection via the airborne mode of transmission. Both SARS-CoV-2 and influenza virions are approximately 0.1 μm in size and can be spherical.^16,17^ Infectious aerosols and droplet nuclei carrying SARS-CoV-2 and influenza are largely spherical, have a material density of approximately 1 g cm^-3^, have a ζ-potential modestly below zero and are polydisperse but, by definition, >0.1 μm based on the size of the virions.^18-20^

For this study, we considered the above parameters and healthcare-relevant experimental conditions to investigate how aerosols penetrate through and leak into N95 FFRs. We chose a polydisperse (0.1 to 1.0 μm) challenge aerosol of spherical Latex polystyrene beads (material density, 1.05 g cm^-3^; ζ-potential < 0, although aerosols were charge neutralized during experimentation for a ζ-potential modestly below zero). Preliminary findings showed that penetration followed a power relationship with aerosol size so that the behavior of aerosols larger than 1.0 μm could be extrapolated using the experimental size range; we did not include aerosols between 1 and 5 μm in this study. For our experimental conditions, we simulated ambient healthcare conditions and its gaseous phase, accounting for the relevant density, dynamic viscosity, breathing rates, ambient temperature, relative humidity (RH), ambient pressure. Further details are included in the Methods section below.

### Methods

#### Implementation of reprocessing methods for N95 FFRs

The seven reprocessing methods assessed in this study include traditional sterilization and decontamination methods in medical settings, emerging ones and processes that have received emergency use authorization (EUA) from the U.S. Federal Drug Administration (FDA).^21-27^ Each reprocessing method, and number of cycles (1, 3, 5 or 10), was evaluated against three models of NIOSH-approved N95 FFRs (3M 1860S, 3M 8210 and 3M 9210, The 3M Company, St. Paul, MN, USA). These models are used widely by healthcare workers (HCWs) and vary in mask design (molded or pleated), strap material (polyisoprene, thermoplastic elastomer and blue polyisoprene for 3M 1860S, 3M 8210 and 3M 9210, respectively) and the presence (3M 1860S) or absence of a colored dye on the exterior surface. All reprocessed N95 FFRs were characterized for leakage or penetration after one day or longer after the last cycle was completed, as described below. Each reprocessing cycle was run using standard parameters or one that have been reported as used for decontamination.^21-27^

For autoclave reprocessing, the N95 FFRs were placed inside of a benchtop autoclave sterilizer (3850E Autoclave, Tuttnauer, Hauppauge, NY, USA), such that no FFR touched another one. For each cycle, they were run under the dry setting (steam time, 30 min) with a 60-min dry time. The N95 FFRs were removed from the autoclave and allowed to sit idly in ambient conditions (30 min) before proceeding.

For vEtOH (70%) reprocessing, we prepared 70% EtOH by mixing the appropriate ratio of ethanol (Sigma-Aldrich, Oakville, ON, Canada) with MilliQ water (18.2 MΩ cm, Milli-Q® IQ 7000 Ultrapure Lab Water System; Millipore Sigma, Etobicoke, ON, Canada). A vapor, vEtOH (70%), was generated via a thin-layer chromatography atomizer (Chemglass Life Sciences, Vineland, NJ, USA) and a fume hood air supply (operated at ~25 psi). For each cycle, the N95 FFRs were covered with vEtOH (70%) and allowed to dry completely under hood ventilation (~1 h) before proceeding.

For forced-air dry heat (100 °C) reprocessing, the N95 FFRs were placed within a benchtop forced air oven (chamber volume, 3.65 ft^3^, VWR® Forced Air Oven; VWR International, Mississauga, ON, Canada), such that no FFR touched another one. For each cycle, the N95 FFRs were heated to 100 °C (ramp time, ~2 min) for 30 min. Afterwards, they were removed from the heat and allowed to cool down to and sit idly at room temperature in ambient conditions (30 min) before proceeding.

For humid heat (75% RH, 75 °C) reprocessing, the N95 FFRs were enclosed within STERIL-PEEL® sterilization pouches (GS Medical Packaging, Inc., Etobicoke, ON, Canada) and placed in a convection heating system with controlled humidity (HCSS74W12, Climate Select Heated Holding Cabinet with Humidity, BevLes Company, Inc., Erie, PA, USA). A humidity gauge (PT2470 Digital Combometer, Exo Terra, Montreal, QC, Canada) was used to ensure that the RH was maintained. For each cycle, the N95 FFRs were heated at 75 °C with 75% RH for 1 h. Afterwards, N95 FFRs were removed from the heat and allowed to cool down in ambient conditions (5 min) before proceeding.

For HPGP reprocessing, the N95 FFRs were enclosed within Tyvek® self-seal sterilization pouches (GS Medical Packaging, Inc., Etobicoke, ON, Canada) and placed in a STERRAD® 100S Sterilizer (Advanced Sterilization Products, Irvine, CA, USA). The N95 FFRs were run through the STERRAD® 100S Long Cycle (59% H_2_O_2_; approximately 72 min per cycle, including venting; 42-50°C; cycle pressure, fluctuated from vacuum to sterilant injection and diffusion to plasma settings with range of 0.3-14.0 Torr). STERRAD® chemical indicator strips (Advanced Sterilization Products, CA, USA) within the sterilization pouches verified exposure during each cycle. The enclosed N95 FFRs were handled after venting.

For HPV reprocessing, the N95 FFRs were enclosed within Vis-U-All^TM^ Low Temperature Sterilization Pouches (STERIS Corporation, OH, USA) and placed in a STERIS V-PRO® maX Low Temperature Sterilization System (STERIS Corporation, OH, USA). Each cycle was run under the non-lumen cycle settings (59% H_2_O_2_, approximately 28 min per cycle, including aeration, 49.3-50.6 °C; cycle pressure, fluctuated from vacuum to sterilant injection settings with 4 pulsations varying from 1-504 Torr). Chemical indicator strips (STERIS Corporation, Ohio, USA) within the sterilization pouches verified exposure during each cycle. The enclosed N95 FFRs were handled after aeration.

For UVGI reprocessing, we constructed an aluminum enclosure containing a SaniRay® RRDHO36-4S High-Output Germicidal Ultraviolet Fixture (Atlantic Ultraviolet Corporation, Hauppauge, NY, USA) with four 254-nm UVC lamps (UV 05-1060-R, Atlantic Ultraviolet Corporation, Hauppauge, NY, USA) mounted in parallel. The enclosure measured 106.68 cm × 60.96 cm × 60.96 cm and was built with an aluminum door to safely introduce and remove samples while containing radiation during operation. A height-adjustable platform was installed and set to 30.48 cm below the lamps for this application. The lamps were warmed up (2 h) to stabilize the UVC irradiance. A UV512C Digital UVC Light Meter (General Tools & Instruments, Secaucus, NJ, USA) was used inside of the enclosure at a fixed position to account for potential fluctuations of UVC irradiation. The UVC irradiance at different areas on the N95 FFRs were mapped using a USB4000 fiber optic spectrometer (Ocean Optics, Dunedin, FL, USA) with a CC-3 Cosine Corrector (Ocean Optics, Dunedin, FL, USA) using 25-scan averages. The results indicated that for the face-side up orientation, the edges of the mask received 57.6% of the irradiance, while the center of the mask received 145.3% of the dose, based on the reference UVC meter. For the face-side down orientation, zones with the lowest irradiance and highest irradiance received 79.4% and 137.3% of the measured reference irradiance, respectively. The N95 FFRs were placed within the UVC enclosure and irradiated face-side up such that all areas on the face-side up orientation received a minimum of ~0.5 J/cm^2^, while being rotated 90° in 30-s intervals to ensure homogeneous dosing. The FFRs were then flipped face-side down and irradiated in the same manner. The process was repeated such that all areas of the N95 FFRs received ~1 J/cm^2^ of UVC or greater. The least exposed areas of the face-up orientation received a UVC dose of 1.010 ± 0.035 J/cm^2^ at an irradiance of 2431 ± 179 μW/cm^2^, while the least exposed areas of the face-down orientation received 1.029 ± 0.039 J/cm^2^ at 3537 ± 199 μW/cm^2^.

#### Characterization of leakage

We characterized leakage via quantitative fit testing. For each pristine N95 FFRs, fit-verified individuals donned and molded an N95 FFRs before assessing leakage. For each reprocessed N95 FFRs, fit-verified individuals (fit factor for pristine masks, 200+) donned and molded an N95 FFRs, doffed it, had it reprocessing using the specified method and number of cycles, and re-donned and molded it to assess leakage. Fit testing was performed using the CSA Z94.4-11 testing standard (PortaCount Respirator Fit Tester 8048, TSI Incorporated, Shoreview, MN, USA), fulfilling OSHA 29CFR 1910.134. Briefly, a sequence of breathing exercises (normal breathing, deep breathing, breathing while turning head side to side, breathing while nodding head up and down, breathing while talking out loud, breathing while bending over and, again, normal breathing) was performed in the proximity of an aerosol generator (Model 8026, TSI

Incorporated; generated aerosols containing NaCl particles, 0.02 μm to >1.0 μm). Since the testing standard and the condensation nuclei counter within the PortaCount instrument exclusively assesses particles between 40 and 70 nm, only particles that leaked through imperfections in facial seal were quantified, rather than those that penetrate through the filter media. These results correspond to leakage due to larger aerosols.^28^

Fit factor (*FF*) is defined by

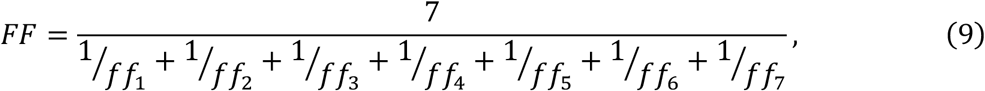

where *ff_i_* is the individual fit score for the *i*-th exercise. Since *FF* is the mean geometric ratio between the concentrations of the test aerosol inside and outside of the N95 FFR (*C_in_*/*C_out_*), leakage was calculated as the inverse (*L* = *FF*^−1^). Within the relevant aerosol size range, leakage is a bulk, size-independent measurement, as leakage occurs through macroscopic imperfection of facial seal. Hence, we took leakage to be a constant value throughout the penetration challenge aerosol range (0.1 to 1.0 μm) when calculating overall transmission. To ensure consistent results (sensitivity, 0.10%), ambient counts were generally maintained above 150 throughout each test. The output value of the testing standard saturates at 200+. Since an output *FF* of 200 corresponds to a leakage of 0.5% and the sensitivity was 0.10%, we considered the limit of detection to be 0.49%.

For reprocessed N95 FFRs, leakage measurements were ensured to exclusively quantify the effect of reprocessing. When re-donning a reprocessed N95 FFR, there is a risk that leakage occurs due to human-based error. To mitigate this issue, fit-verified individuals re-donned reprocessed N95 FFRs while viewing the live *FF*. The live *FF* was maximized before testing, meaning that increases in leakage were due to the effects of a reprocessing method and number of cycles.

HCWs widely exhibit one or more behaviors of improper wear for N95 FFRs, such as pinching the nose clip while molding the mask.^29,30^ We simulated this common compliance issue. Fit-verified individuals donned and molded a pristine N95 FFR without viewing the live *FF*. While doing so, they molded the nose clip outward in, rather than the recommended inward out, thereby pinching the nose clip and creating a relatively sharp bend at the apex of the nose clip. Leakage was then quantified for these masks.

#### Characterization of penetration and pressure differential

Penetration experiments were performed at SGS-IBR Laboratories (Grass Lakes, MI, USA). These aerosols and experimental conditions simulated those found in healthcare settings and for moderate HCW breathing through N95 FFRs.^31,32^ To standardize experimentation, penetration measurements were conducted according to particle filtration efficiency measurements for ASTM F2299 and ASTM F2100.^33,34^ As previously introduced, we characterized penetration using a polydisperse aerosol of negatively charged spherical Latex polystyrene beads. Briefly, we mixed monodisperse aqueous suspensions of Latex polystyrene microspheres for a polydisperse distribution of challenge particles (0.1 μm to 1 μm; material density, 1.05 g cm^-3^ at 20 °C). Filtered and dried air was passed through a nebulizer to produce an aerosol containing the suspended Latex microspheres. The aerosol was passed through a charge neutralizer, leading to a ζ-potential modestly below 0, and mixed and diluted with additional preconditioned air to produce the challenge aerosol to be used in the test. N95 FFRs were tested previously for leakage and contained fit test sampling probes (TSI Incorporated, Shoreview, MN, USA). Leftover sample probes were sealed with hot glue, and control N95 FFRs with sealed probes were indistinguishable from control ones without them based on penetration and pressure differential measurements. N95 FFRs were attached to a filter holder and placed between inflow and outflow tubes. The aerosol was fed (1.0 scfm) through the FFRs, and penetration was obtained using two particle counters (Lasair® III 110 Airborne Particle Counter, Particle Measuring Systems®, a Spectris company Boulder, CO, USA) connected to the feed stream and filtrate. Penetration was measured within six size channels (0.1 to 0.15 μm, 0.15 to 0.20 μm, 0.20 to 0.25 μm, 0.25 to 0.30 μm, 0.3 to 0.5 μm and 0.5 to 1.0 μm). For power regression (described in the section below), we took the measured penetration within each channel to be the middle of its size channel (0.125 μm, 0.175 μm, 0.225 μm, 0.275 μm, 0.4 μm and 0.75 μm). This was justified based on high coefficients of determination (*R^2^*) throughout the samples in this study and because expectation values for penetration were conservative estimates, with expected penetration typically being slightly greater than the experimental values. Pressure differential (DHII-007, Dwyer Instruments International, Michigan City, IN, USA), air flow (M-50SLPM-D/5M, Alicat Scientific, Tucson, AZ, USA), temperature and humidity (HMT330 Humidity and Temperature Meter, Vaisala, Helsinki, Finland) and barometric pressure (PTU200 Transmitter, Vaisala, Helsinki, Finland) were also characterized in the experimental apparatus. Pressure differential was measured for greater mechanistic insight into how reprocessing affected penetration.^26^ Throughout the penetration experiments, the temperature, relative humidity and barometric pressure were measured to be 20.9 ± 0.52 °C, 48.5 ± 3.70 % and 723.6 ± 2.72 mmHg, respectively. Note that higher RH values (e.g., 80%, like the preconditioning stage for the NIOSH test) is not expected to affect penetration measurements.^35^

#### Statistical analyses

For power regression, the statistical model had the traditional nonlinear form,

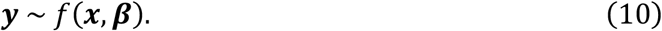

We fitted the discrete aerosol penetration measurements for each sample according to the model

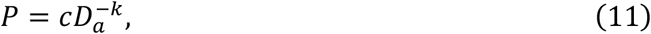

where *P* is the expected penetration, *c* is the scaling constant, *D_a_* is the aerosol size and *k* is the determined power law exponent. We used the least squares estimator

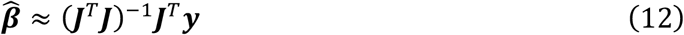

and assumed that the model could be approximated using a first-order Taylor series

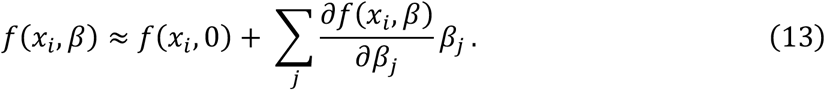

Expectation curves and confidence bands were generated using these approximations at a sufficient number of intervals (*n* = 50) throughout the axes spans and by linearly connecting them. We applied power regression to the discrete penetration data from individual samples. A strong cumulative coefficient of determination (*R*^2^, 0.94 ± 0.041, mean ± S.E.M.) across all samples in included in this study suggested that this power law was a good model for the relationship between penetration and aerosol size.

We performed equivalence testing to compare the likeness of the aerosol transmission characteristics of pristine N95 FFRs and reprocessed ones. We applied the conventional two-one-sided t-test procedure^36^ and took the ratio between overall transmission for all N95 FFRs, including across models, that were reprocessed using a certain method and number of cycles and all properly worn, pristine N95 FFRs. Since equivalence bounds are not standardized in this field, we used the U.S. FDA’s standard bounds for bioequivalence (upper equivalence bound = 1.25, lower equivalence bound = 0.80, based on the geometric mean ratio).^36^ N95 FFRs reprocessed using each method for one cycle were tested first. For some reprocessing methods, our results showed rejection of the null hypothesis (*P* < 0.01 and *P* < 0.001). If these reprocessed masks accepted the null hypothesis (*P* > 0.05), we did not perform equivalence testing for higher cycle numbers. N95 FFRs reprocessed using three methods passed at one cycle, but none did at three cycles.

### Limitations of this study

#### Composition of matter for N95 FFRs

The three models of N95 FFRs used in this study span a range of mask designs and constituent materials and are widely used by HCWs. Nevertheless, there are other models and brands used by HCWs which may differ in the composition of matter. Of note, the three 3M models studied use an electret for the filter media that can restore static charge over time.^37^ Since we found that reprocessing mainly increased aerosol penetration by degrading filter charge, our results may underestimate the impact of certain reprocessing methods on aerosol penetration for N95 brands and models without electret properties. The N95 FFRs used in this study included those from several batches manufactured years apart. Differences arising from batch manufacturing may be encapsulated in this study. Batch manufacturing may also skew the transmission characteristics of an N95 FFR.

#### Relevance to reprocessing N95 FFRs in healthcare settings

This study focuses on the direct assessment of aerosol transmission for pristine, improperly worn pristine and reprocessed, but properly worn, N95 FFRs. Our results help to understand the exposure risk for HCWs performing AGMPs or near other sources of infectious aerosols. They also help to understand the implications of reprocessing. We did not, however, investigate the effects of HCW wear, especially when extended-use guidelines are implemented. In addition, the field does not currently understand the extensiveness and impact of extended-use guidelines on noncompliance in wear. These, and additional contributions, may adversely affect the aerosol transmission characteristics of N95 FFRs in healthcare settings. For example, we showed that at one cycle of forced-air dry heat, HPGP or HPV reprocessed N95 FFRs were statistically equivalent to pristine ones in terms of aerosol transmission. This result does not account for the effects of extended wear, which may affect performance. In addition, for proper experimental design, we evaluated leakage on fit-verified individuals who had an optimal fit factor (200+). Since quantitative fit testing considers a fit factor of 100 to be a pass, some institutions may allow HCWs to wear N95 FFRs that do not fit optimally, increasing leakage by a predictable amount. From one perspective, the results in this study can be taken as approximate better-case scenarios (i.e., upper bounds), especially for greater reprocessing cycle numbers. For proper clinical implementation of reprocessing for N95 FFRs, the effects of the aforementioned contributions on filtration performance should be studied.

## Supplementary figures

**Figure S1.**
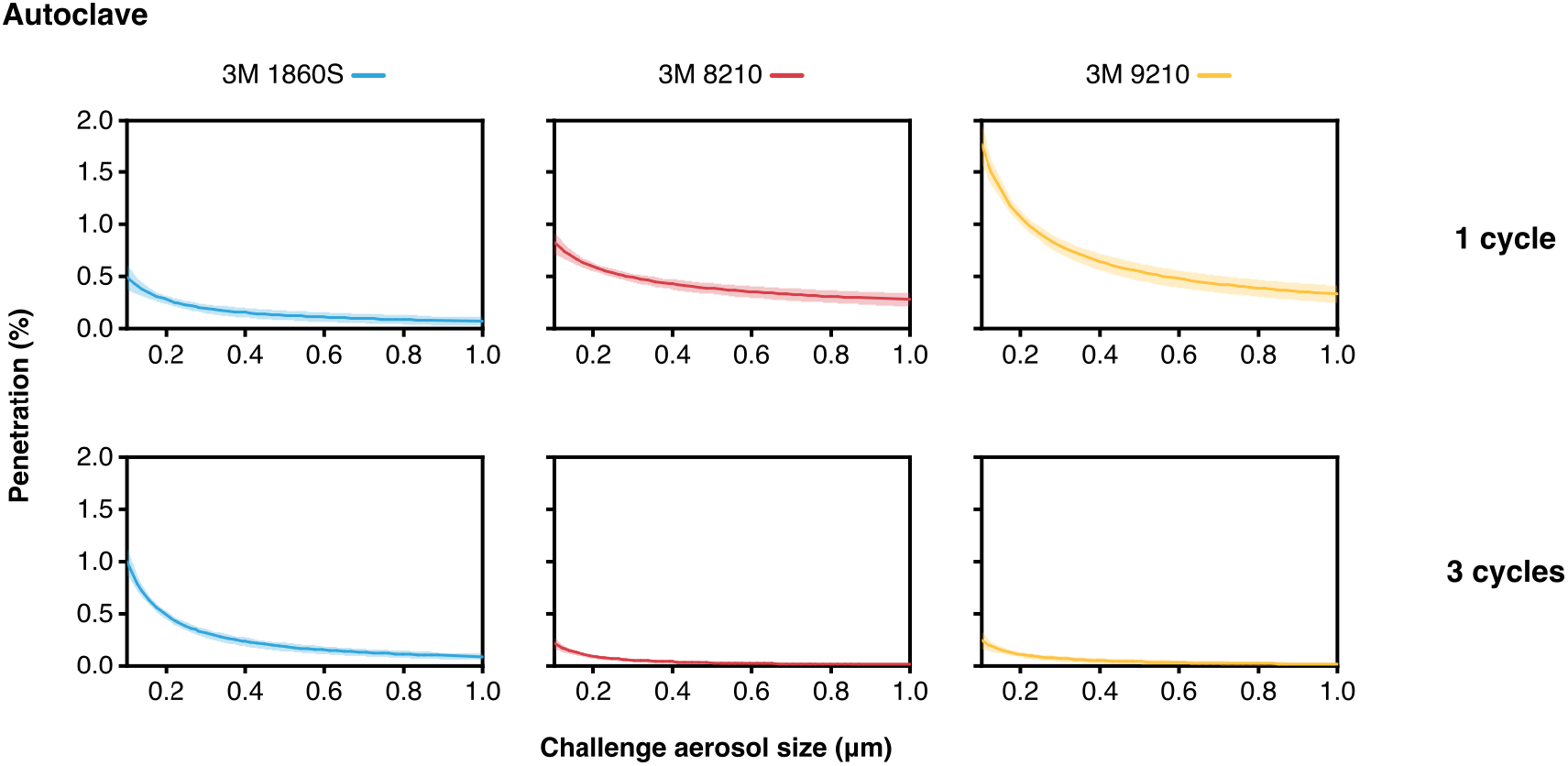
Penetration plots for N95 FFRs that were reprocessed via autoclave for 1 or 3 cycles. Samples were not reprocessed using autoclave for 5 or 10 cycles. Curves and bands depict the expectation line and its 95% confidence band, respectively, from power regression for individual samples.

**Figure S2.**
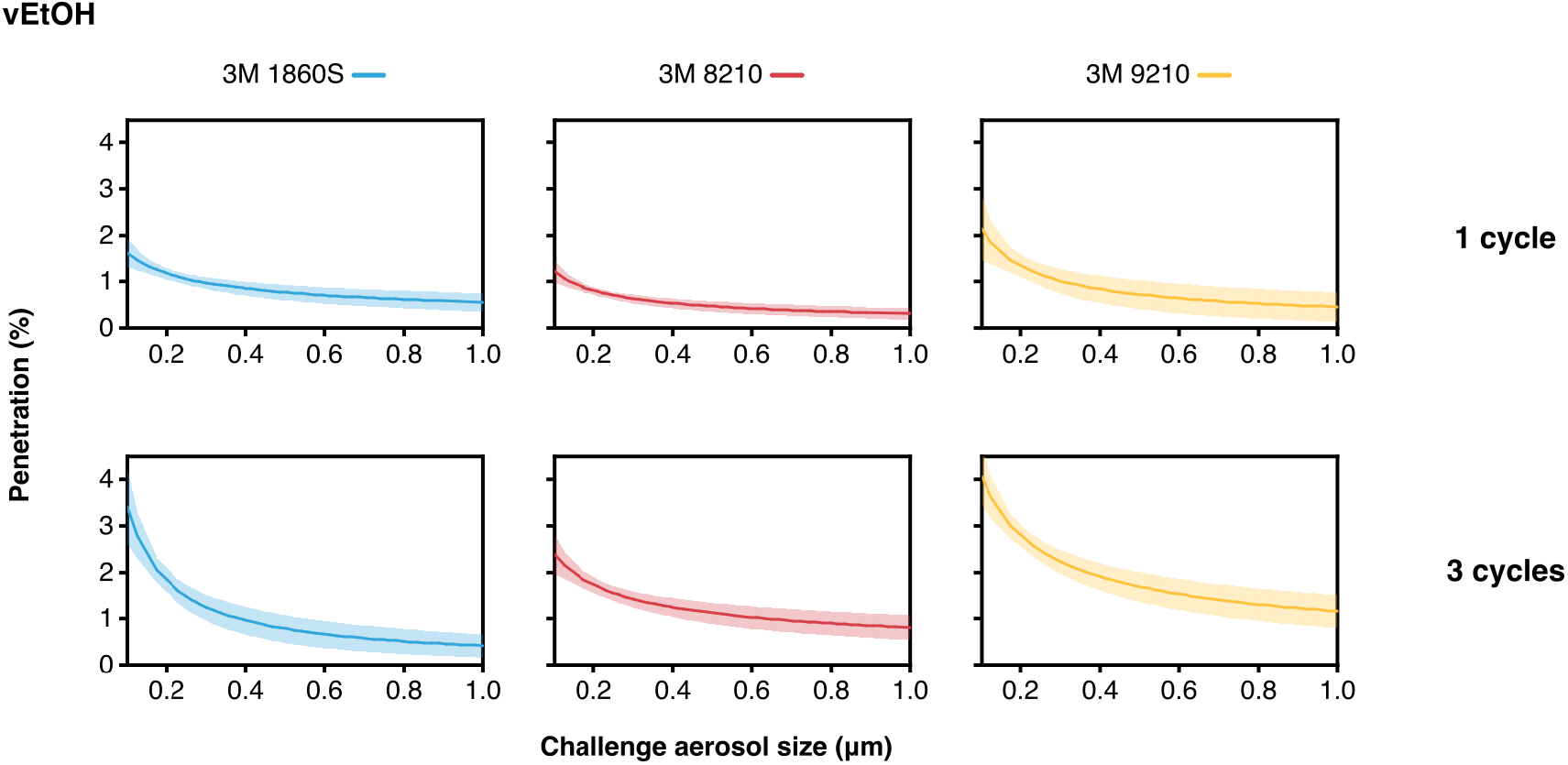
Penetration plots for N95 FFRs that were reprocessed via vEtOH (70%) for 1 or 3 cycles. Samples were not reprocessed using autoclave for 5 or 10 cycles. Curves and bands depict the expectation line and its 95% confidence band, respectively, from power regression for individual samples.

**Figure S3.**
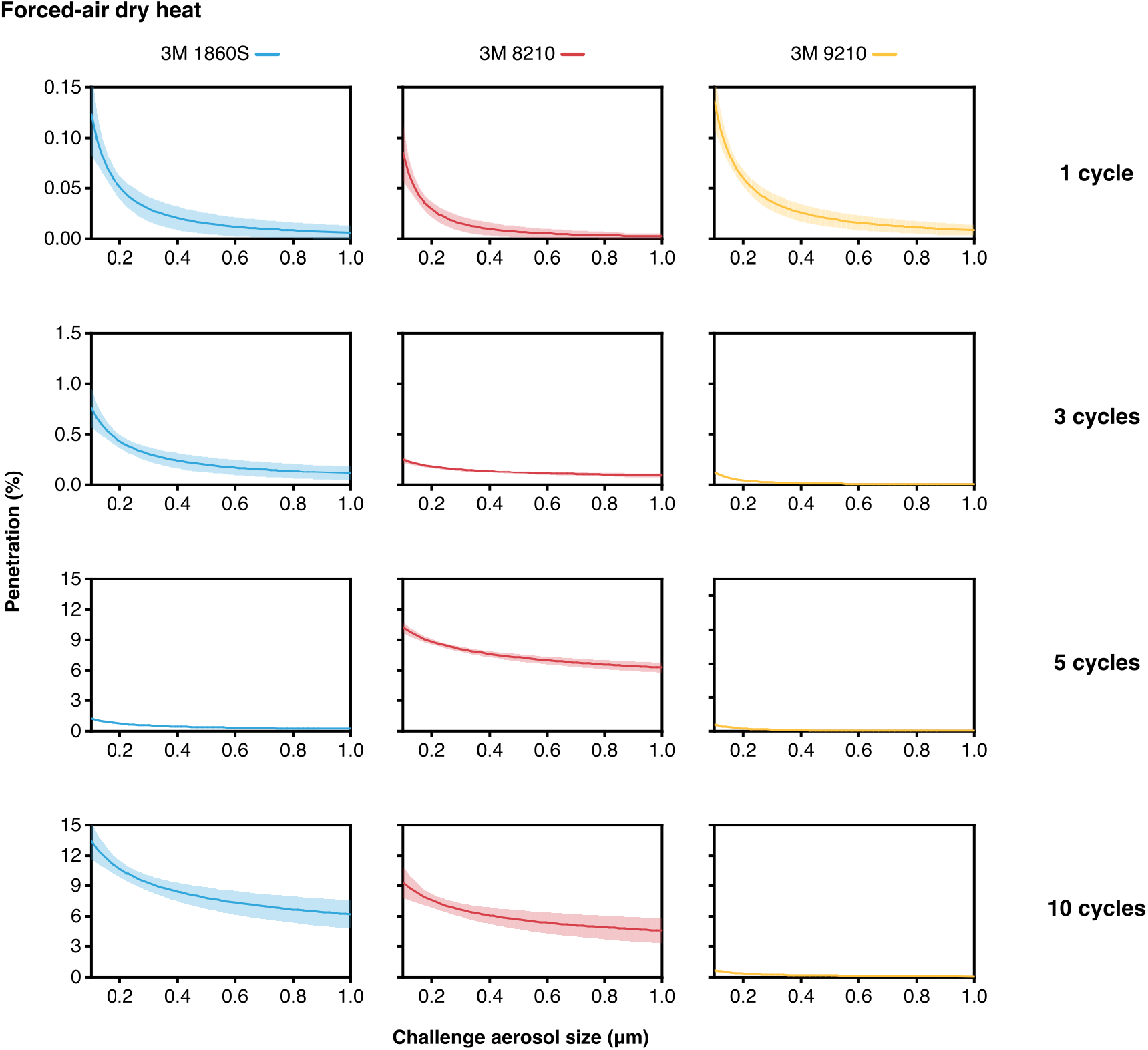
Penetration plots for N95 FFRs that were reprocessed via forced-air dry heat (100 °C) for 1, 3, 5 or 10 cycles. Curves and bands depict the expectation line and its 95% confidence band, respectively, from power regression for individual samples.

**Figure S4.**
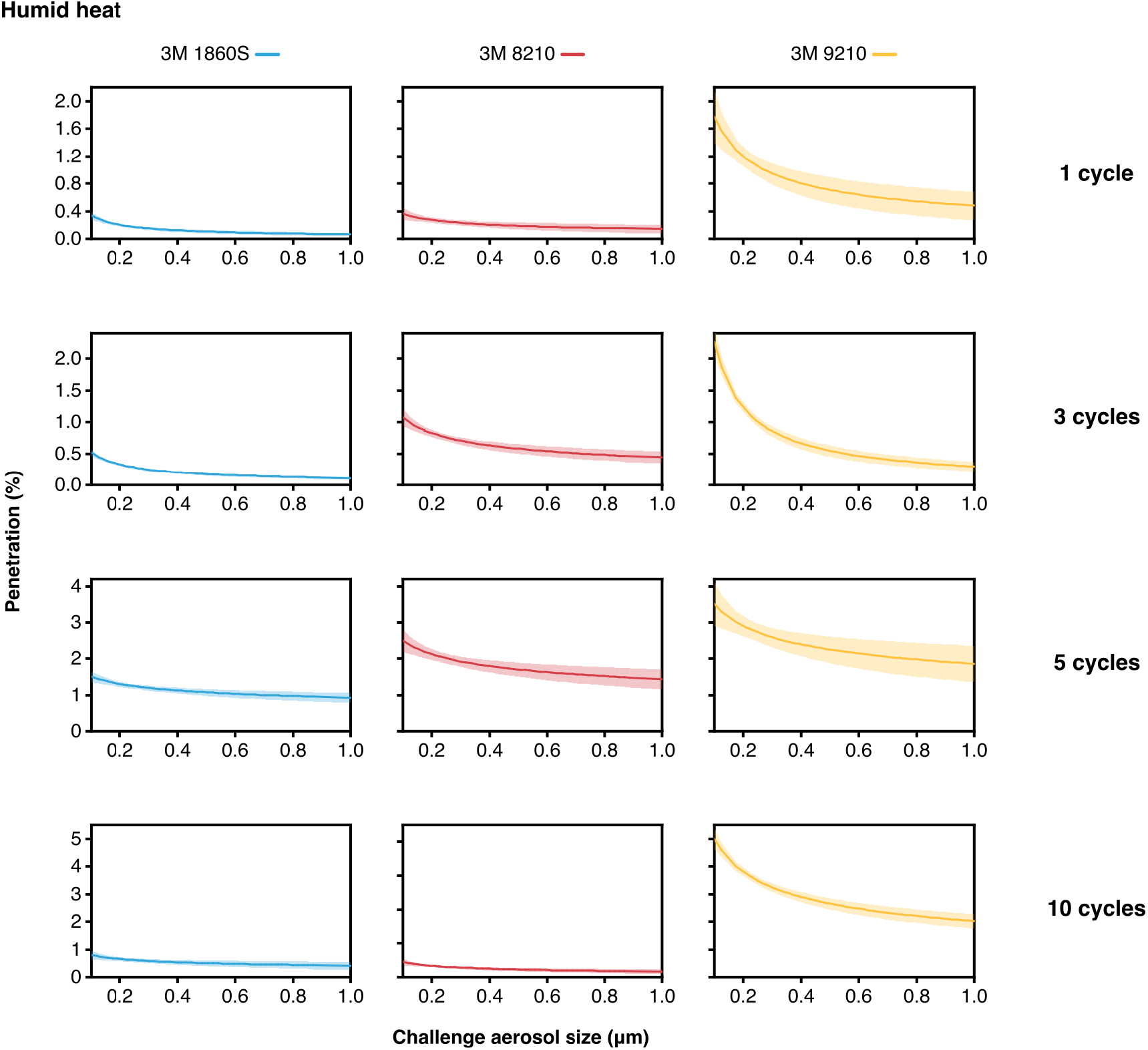
Penetration plots for N95 FFRs that were reprocessed via humid heat (75% RH, 75°C) for 1, 3, 5 or 10 cycles. Curves and bands depict the expectation line and its 95% confidence band, respectively, from power regression for individual samples.

**Figure S5.**
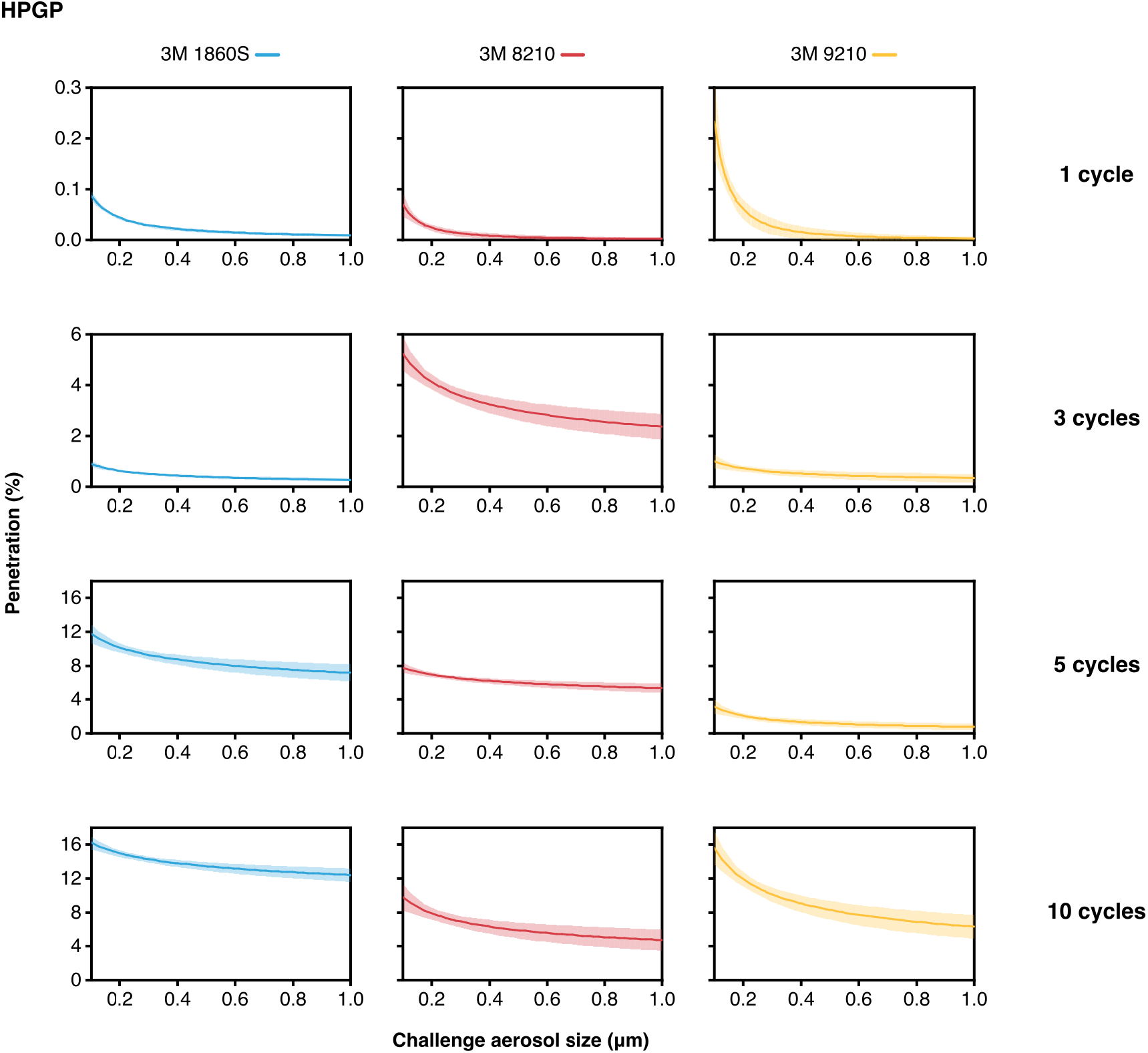
Penetration plots for N95 FFRs that were reprocessed via HPGP (STERRAD® 100S) for 1, 3, 5 or 10 cycles. Curves and bands depict the expectation line and its 95% confidence band, respectively, from power regression for individual samples.

**Figure S6.**
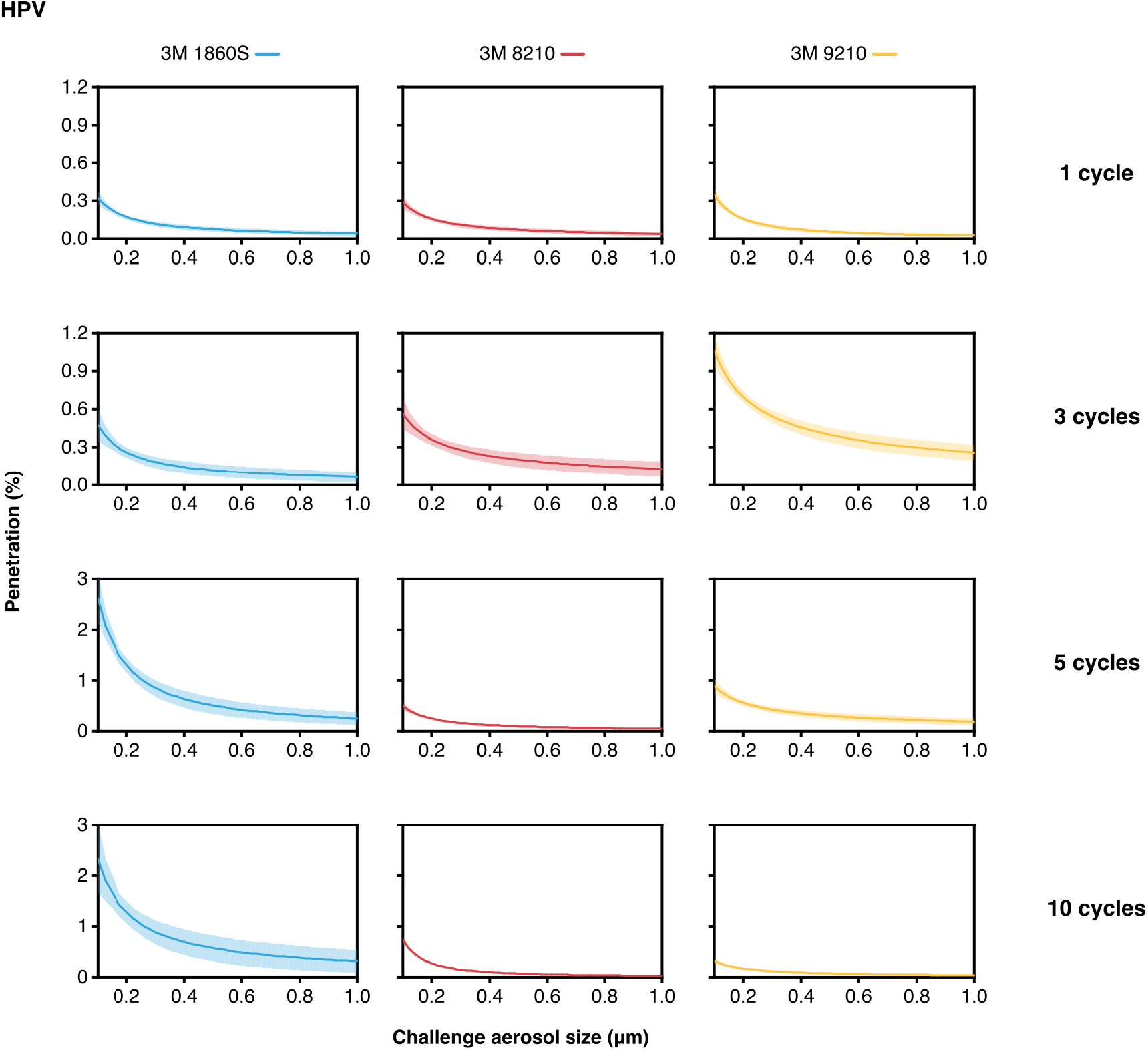
Penetration plots for N95 FFRs that were reprocessed via HPV (STERIS V-PRO®) for 1, 3, 5 or 10 cycles. Curves and bands depict the expectation line and its 95% confidence band, respectively, from power regression for individual samples.

**Figure S7.**
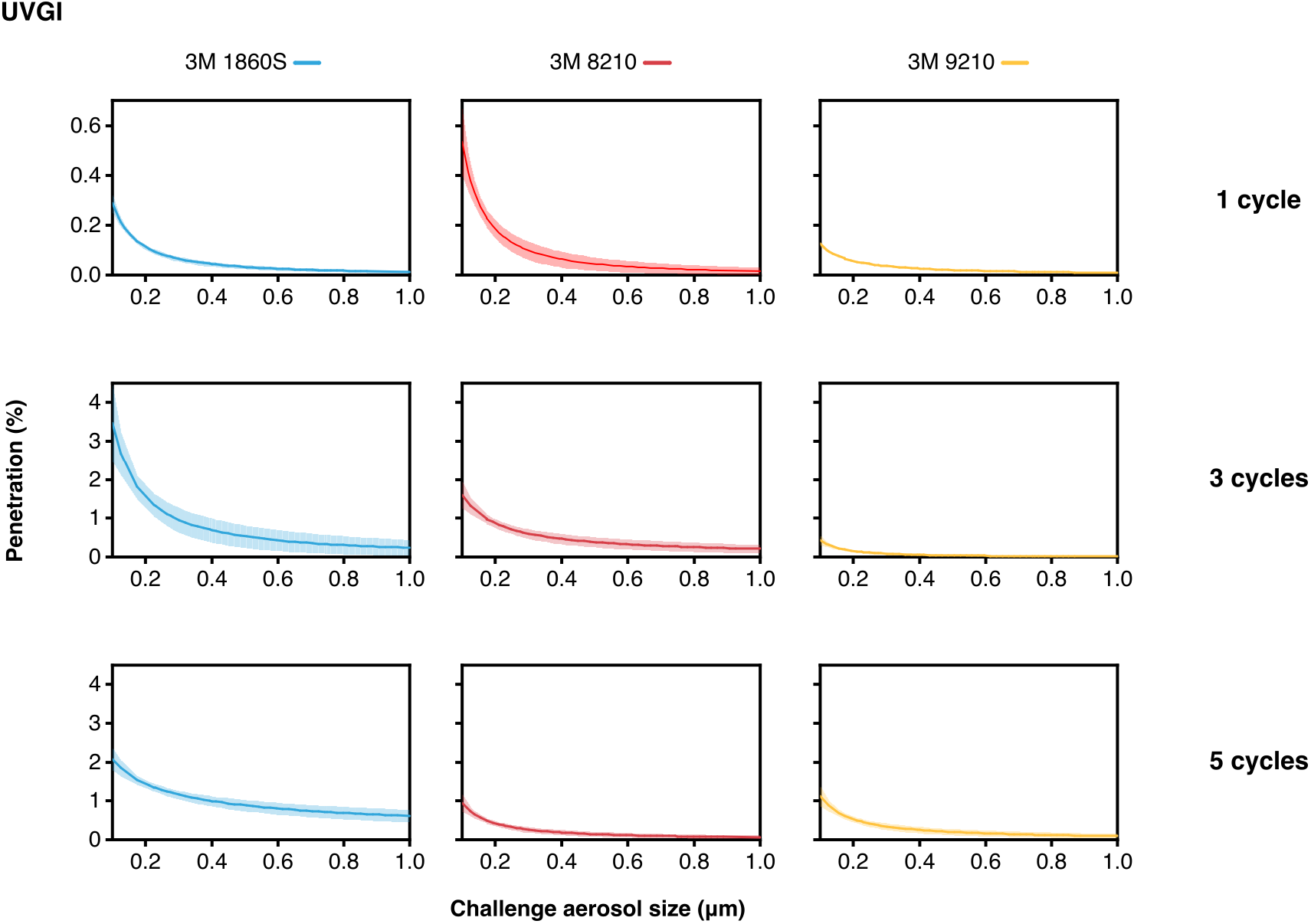
Penetration plots for N95 FFRs that were reprocessed via UVGI for 1, 3 or 5 cycles. Samples were not reprocessed using autoclave for 10 cycles. Curves and bands depict the expectation line and its 95% confidence band, respectively, from power regression for individual samples.

**Figure S8.**
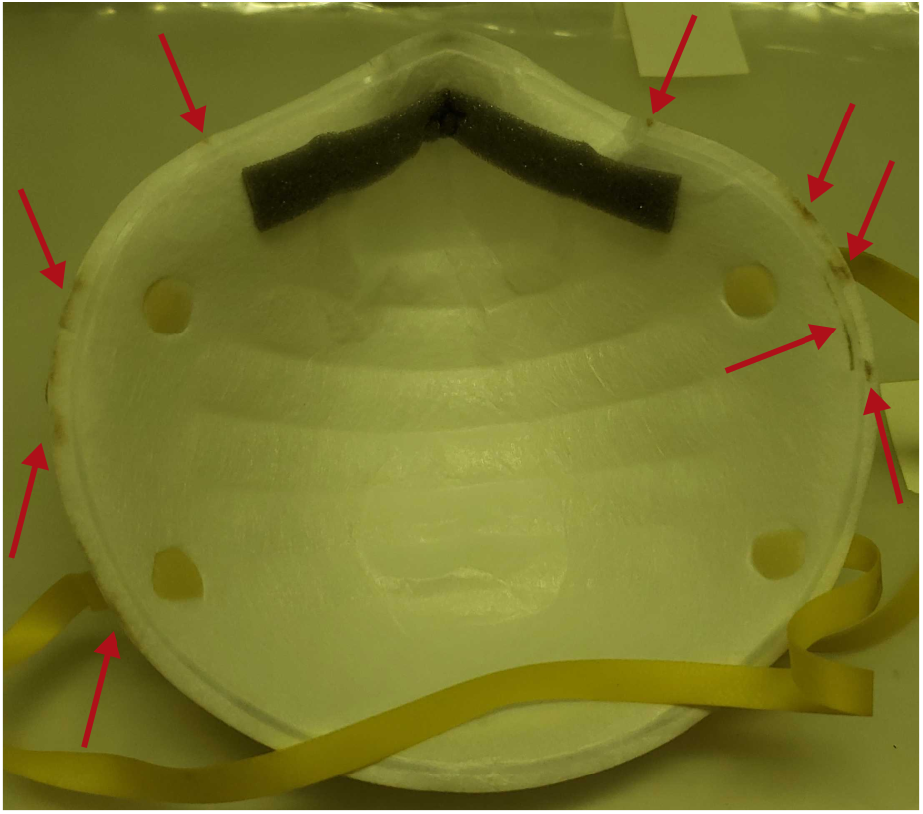
UVGI reprocessing can potentially induce dose-dependent photochemical damage to N95 FFRs. Image of an N95 FFR (3M 8210) that has undergone 3 reprocessing cycles and displays slight damage, as depicted by the red arrows.

**Figure S9.**
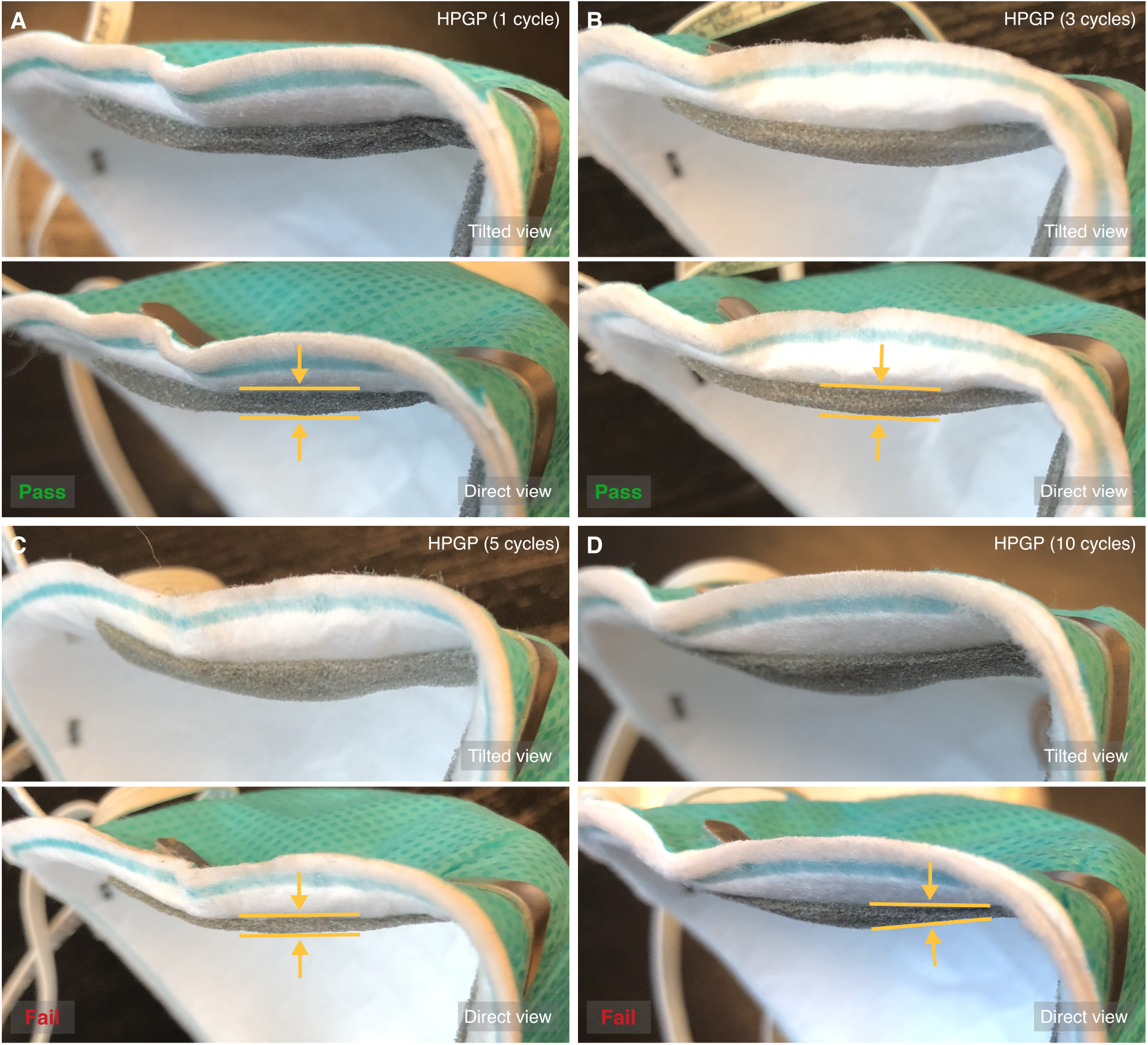
HPGP (STERRAD® 100S) reprocessing degrades the polyurethane nose foam of N95 FFRs. Tilted (top) and direct (bottom) images of the nose foam of 3M 1860S FFRs after 1 cycle (A), 3 cycles (B), 5 cycles (C) and 10 cycles (D) of HPGP reprocessing. Pass or fail refers to the results from quantitative fit testing. At 5 and 10 cycles, nose foams felt brittle. The yellow markings denote the thickness of each nose foam. The HPGP and HPV cycles run are essentially similar (e.g., H2O2 concentration and experimental conditions) except for the plasma phase of HPGP. Since HPV did not induce nose foam degradation, these results suggest the hydroxyl and hydroperoxyl radicals from the plasma oxidize the polyurethane nose foams across N95 models.

**Figure S10.**
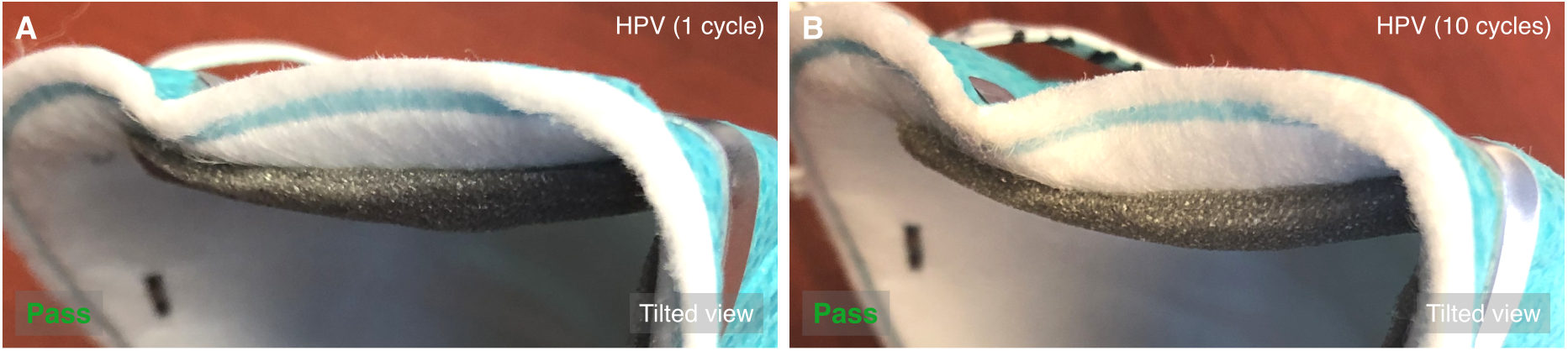
HPV (STERIS V-PRO® maX) reprocessing maintains the polyurethane nose foam of N95 FFRs for at least 10 cycles. Tilted images of the nose foam of 3M 1860S FFRs after 1 cycle (A) and 10 cycles (B) of HPV reprocessing. Pass refers to the results from quantitative fit testing. In addition, there was no noticeable impact on the feel of the nose foam up to 10 cycles.

**Figure S11.**
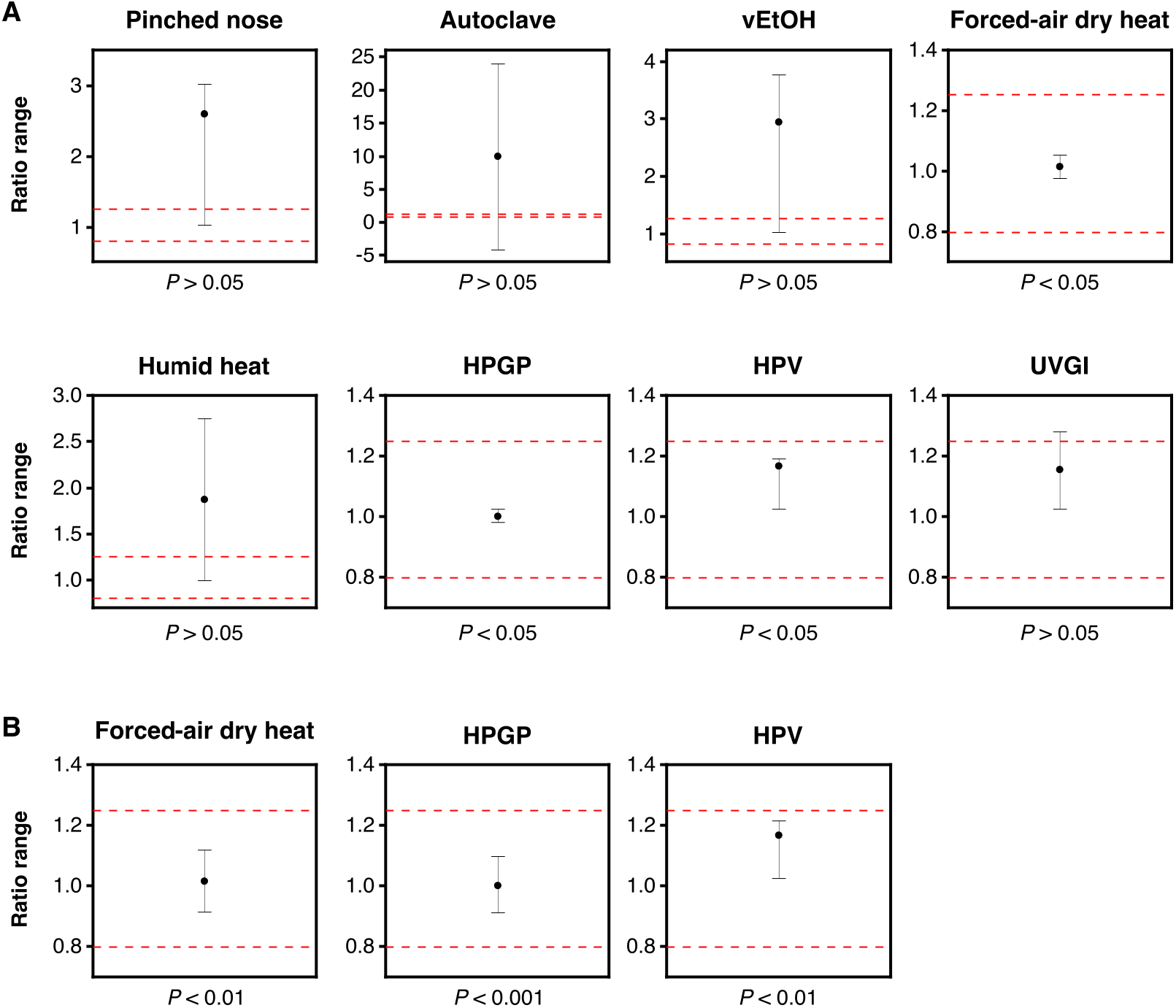
Equivalence testing compares the overall transmission of pristine N95 FFRs with that of improperly worn N95 FFRs (pinched nose clip) or those that have been reprocessed for 1 cycle via the seven methods (geometric mean ratio, upper equivalence bound = 1.25, lower equivalence bound = 0.80) with α = 0.05 (A) or α = 0.01 or α = 0.001 (B). The dots and I bars represent the geometric mean ratios and their 100(1 - 2α)% confidence intervals, respectively. The red dashed lines represent the upper and lower equivalence bounds. The f-value inequalities are reported below each plot.

**Figure S12.**
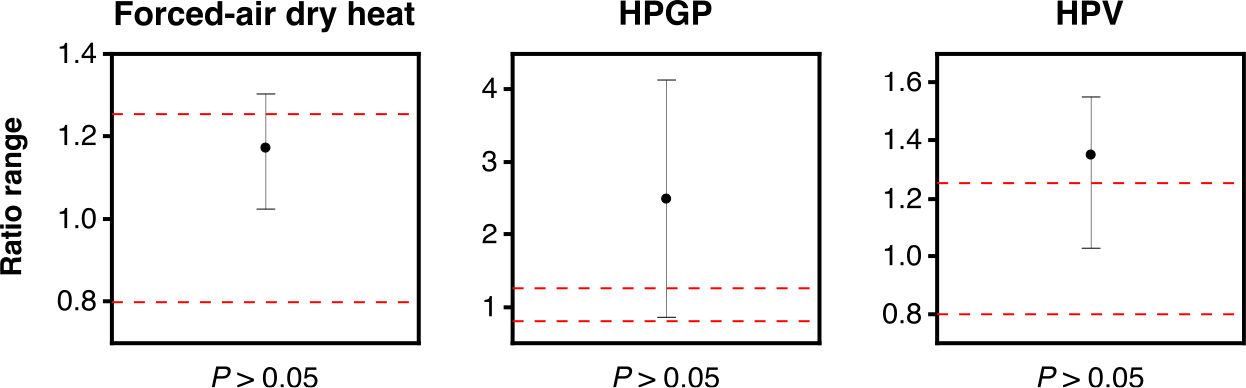
Equivalence testing compares the overall transmission of pristine N95 FFRs with reprocessed ones for 3 cycles via forced-air dry heat (100 °C), HPGP (STERRAD® 100S) or HPV (STERIS V-PRO® maX) (geometric mean ratio, upper equivalence bound = 1.25, lower equivalence bound = 0.80, α = 0.05). The dots and I bars represent the geometric mean ratios and their 100(1 - 2α)% confidence intervals, respectively. The red dashed lines represent the upper and lower equivalence bounds. The *P*-value inequalities are reported below each plot.

**Figure S13.**
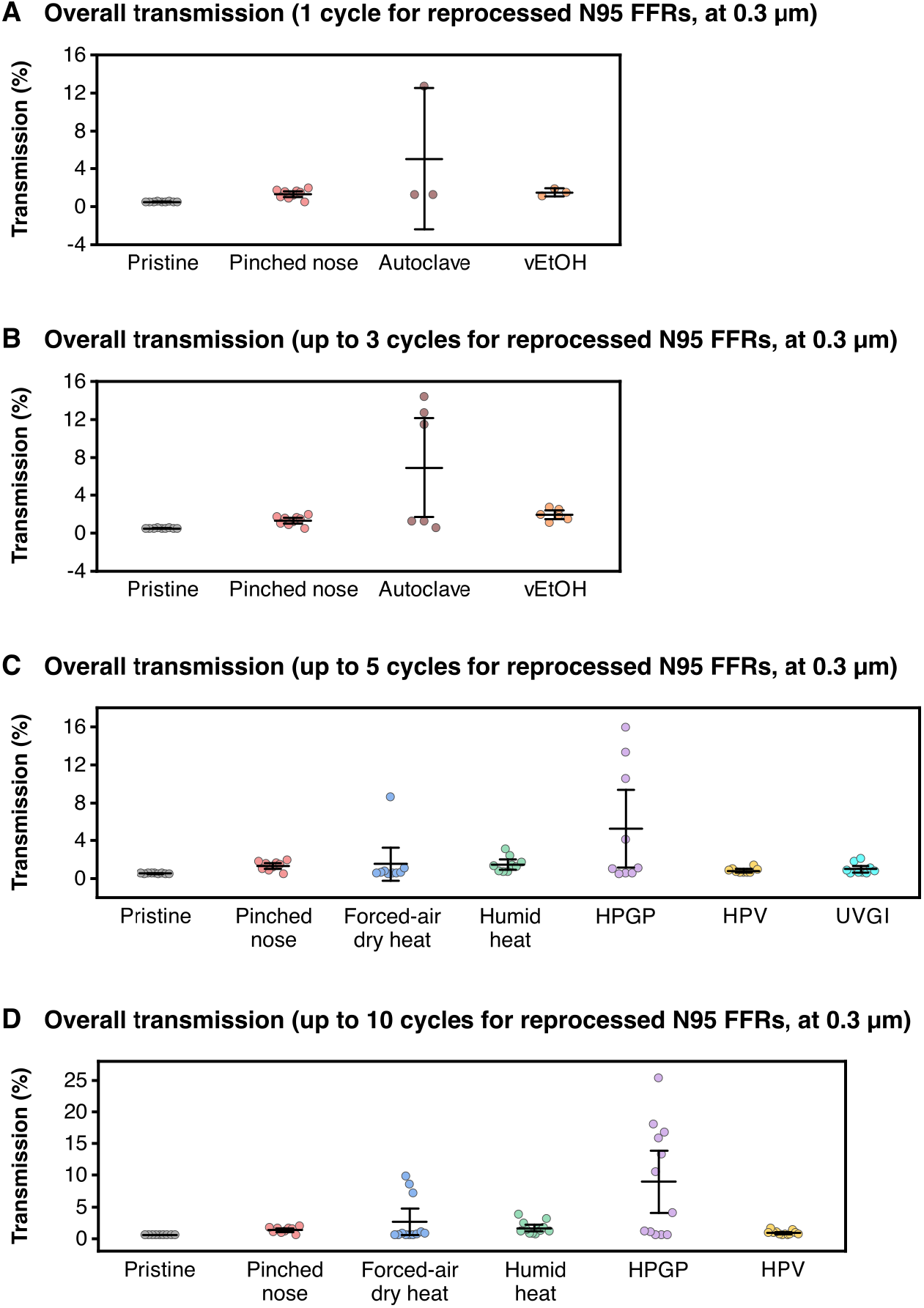
Comparison of the overall transmission for pristine N95 FFRs, improperly worn pristine N95 FFRs (pinched nose clip) and those that have been reprocessed for 1 (A), 3 (B), 5 (C) or 10 (D) cycles. Individual data points represent the expectation values from power regressions at an aerosol size of 0.3 μm. Data included in the main body (Fig. 1) are excluded in this supplementary figure. A reprocessing method was excluded in (C) (autoclave and vEtOH) and (D) (UVGI) if it was not run for the respective number of cycles. The middle bars and I bars represent the estimate mean and its 95% confidence interval, respectively.

## Supplementary table

**Table S1.** Summary of the aerosol transmission characteristics of pristine and reprocessed N95 FFRs.

| Reprocessing method | Model of N95 FFR evaluated | Number of reprocessing cycles | Leakage (%)<sup>†</sup> | Expected penetration* (%; 0.3 µm) | 95% confidence interval* (%) | R<sup>2</sup> * | Pressure differential (mmH<sub>2</sub>O) |
| --- | --- | --- | --- | --- | --- | --- | --- |
| Pristine (not reprocessed) | 3M 1860S | 0 | ≤0.49 ± 0 | 0.024 | (0.013, 0.035) | 0.94 | 3.6 |
|  | 3M 8210 | 0 | ≤0.49 ± 0 | 0.025 | (0.012, 0.037) | 0.97 | 2.1 |
|  | 3M 9210 | 0 | ≤0.49 ± 0 | 0.019 | (0.013, 0.024) | 0.97 | 2.8 |
| Autoclave | 3M 1860S | 1 | 12.50 | 0.195 | (0.151, 0.238) | 0.94 | 4.6 |
|  |  | 3 | 11.11 | 0.315 | (0.273, 0.356) | 0.99 | 4.3 |
|  | 3M 8210 | 1 | 0.74 | 0.488 | (0.446, 0.529) | 0.96 | 3.3 |
|  |  | 3 | 14.29 | 0.057 | (0.040, 0.074) | 0.96 | 3.3 |
|  | 3M 9210 | 1 | ≤0.49 | 0.789 | (0.717, 0.860) | 0.99 | 3.3 |
|  |  | 3 | ≤0.49 | 0.071 | (0.043, 0.098) | 0.93 | 3.3 |
| vEtOH (70%) | 3M 1860S | 1 | 0.93 | 0.972 | (0.845, 1.099) | 0.92 | 3.6 |
|  |  | 3 | 1.22 | 1.246 | (0.963, 1.528) | 0.95 | 4.3 |
|  | 3M 8210 | 1 | ≤0.49 | 0.633 | (0.540, 0.726) | 0.93 | 3.3 |
|  |  | 3 | 0.52 | 1.424 | (1.251, 1.598) | 0.92 | 3.3 |
|  | 3M 9210 | 1 | ≤0.49 | 1.014 | (0.750, 1.278) | 0.87 | 3.8 |
|  |  | 3 | ≤0.49 | 2.228 | (1.974, 2.481) | 0.95 | 3.3 |
| Forced-air dry heat (100 °C) | 3M 1860S | 1 | 0.51 | 0.029 | (0.017, 0.042) | 0.94 | 4.6 |
|  |  | 3 | ≤0.49 | 0.310 | (0.239, 0.381) | 0.93 | 4.6 |
|  |  | 5 | ≤0.49 | 0.562 | (0.531, 0.594) | 0.99 | 4.6 |
|  |  | 10 | 0.57 | 9.259 | (8.478, 10.040) | 0.93 | 4.3 |
|  | 3M 8210 | 1 | ≤0.49 | 0.010 | (0.003, 0.016) | 0.95 | 4.1 |
|  |  | 3 | ≤0.49 | 0.156 | (0.145, 0.167) | 0.97 | 3.6 |
|  |  | 5 | ≤0.49 | 8.107 | (7.862, 8.352) | 0.97 | 3.6 |
|  |  | 10 | ≤0.49 | 6.638 | (5.955, 7.322) | 0.87 | 3.3 |
|  | 3M 9210 | 1 | ≤0.49 | 0.036 | (0.027, 0.045) | 0.97 | 3.3 |
|  |  | 3 | 0.55 | 0.024 | (0.018, 0.030) | 0.99 | 3.6 |
|  |  | 5 | ≤0.49 | 0.046 | (0.028, 0.065) | 0.95 | 3.3 |
|  |  | 10 | 0.50 | 0.265 | (0.199, 0.331) | 0.93 | 3.8 |
| Humid heat (75% RH, 75 °C) | 3M 1860S | 1 | 0.55 | 0.151 | (0.129, 0.173) | 0.96 | 4.1 |
|  |  | 3 | 0.52 | 0.244 | (0.225, 0.263) | 0.99 | 4.1 |
|  |  | 5 | 0.51 | 1.195 | (1.127, 1.263) | 0.91 | 4.3 |
|  |  | 10 | 0.60 | 0.584 | (0.510, 0.658) | 0.80 | 4.3 |
|  | 3M 8210 | 1 | 0.51 | 0.231 | (0.192, 0.270) | 0.84 | 3.0 |
|  |  | 3 | 0.52 | 0.700 | (0.643, 0.758) | 0.95 | 3.0 |
|  |  | 5 | ≤0.49 | 1.924 | (1.783, 2.065) | 0.89 | 3.6 |
|  |  | 10 | 0.83 | 0.280 | (0.240, 0.320) | 0.89 | 3.3 |
|  | 3M 9210 | 1 | ≤0.49 | 0.945 | (0.793, 1.098) | 0.93 | 3.6 |
|  |  | 3 | ≤0.49 | 0.849 | (0.764, 0.934) | 0.99 | 4.3 |
|  |  | 5 | ≤0.49 | 2.591 | (2.325, 2.858) | 0.83 | 3.3 |
|  |  | 10 | 0.51 | 3.239 | (3.084, 3.393) | 0.98 | 2.8 |
| HPGP (STERRAD® 100S) | 3M 1860S | 1 | ≤0.49 | 0.029 | (0.026, 0.032) | 0.99 | 4.3 |
|  |  | 3 | ≤0.49 | 0.504 | (0.457, 0.551) | 0.97 | 4.3 |
|  |  | 5 | 4.00 | 9.271 | (8.762, 9.780) | 0.92 | 4.3 |
|  |  | 10 | 11.11 | 14.274 | (13.914, 14.633) | 0.94 | 4.1 |
|  | 3M 8210 | 1 | ≤0.49 | 0.013 | (0.007, 0.019) | 0.94 | 3.0 |
|  |  | 3 | ≤0.49 | 3.576 | (3.290, 3.862) | 0.94 | 3.3 |
|  |  | 5 | 4.00 | 6.494 | (6.230, 6.757) | 0.92 | 3.0 |
|  |  | 10 | 11.11 | 6.914 | (6.221, 7.606) | 0.89 | 3.3 |
|  | 3M 9210 | 1 | ≤0.49 | 0.027 | (0.011, 0.043) | 0.96 | 3.8 |
|  |  | 3 | ≤0.49 | 0.589 | (0.473, 0.706) | 0.84 | 3.8 |
|  |  | 5 | 14.29 | 1.607 | (1.291, 1.924) | 0.91 | 3.0 |
|  |  | 10 | 6.67 | 10.108 | (9.271, 10.945) | 0.95 | 4.1 |
| HPV<br>(STERIS V-Pro® maX) | 3M 1860S | 1 | ≤0.49 | 0.118 | (0.098, 0.137) | 0.97 | 4.6 |
|  |  | 3 | ≤0.49 | 0.182 | (0.137, 0.226) | 0.94 | 4.8 |
|  |  | 5 | 0.51 | 0.849 | (0.686, 1.013) | 0.97 | 4.8 |
|  |  | 10 | 0.58 | 0.887 | (0.651, 1.123) | 0.93 | 4.3 |
|  | 3M 8210 | 1 | ≤0.49 | 0.109 | (0.092, 0.126) | 0.97 | 4.3 |
|  |  | 3 | ≤0.49 | 0.163 | (0.137, 0.189) | 0.93 | 3.8 |
|  |  | 5 | 0.57 | 0.277 | (0.230, 0.324) | 0.98 | 3.6 |
|  |  | 10 | 0.52 | 0.149 | (0.129, 0.169) | 0.99 | 3.8 |
|  | 3M 9210 | 1 | ≤0.49 | 0.098 | (0.081, 0.115) | 0.98 | 4.6 |
|  |  | 3 | ≤0.49 | 0.541 | (0.492, 0.589) | 0.98 | 4.3 |
|  |  | 5 | ≤0.49 | 0.424 | (0.364, 0.484) | 0.96 | 4.3 |
|  |  | 10 | ≤0.49 | 0.378 | (0.338, 0.418) | 0.98 | 4.3 |
| UVGI | 3M 1860S | 1 | 0.55 | 0.064 | (0.054, 0.074) | 0.99 | 4.6 |
|  |  | 3 | 0.81 | 0.956 | (0.639, 1.273) | 0.94 | 4.6 |
|  |  | 5 | 0.96 | 1.121 | (1.003, 1.238) | 0.96 | 4.6 |
|  | 3M 8210 | 1 | 0.54 | 0.097 | (0.061, 0.134) | 0.97 | 3.6 |
|  |  | 3 | ≤0.49 | 0.601 | (0.477, 0.726) | 0.96 | 3.8 |
|  |  | 5 | 0.51 | 0.258 | (0.186, 0.330) | 0.96 | 3.6 |
|  | 3M 9210 | 1 | ≤0.49 | 0.035 | (0.032, 0.039) | 0.99 | 3.0 |
|  |  | 3 | ≤0.49 | 0.083 | (0.050, 0.116) | 0.96 | 3.3 |
|  |  | 5 | ≤0.49 | 0.340 | (0.256, 0.424) | 0.97 | 4.6 |

†The limit of detection for leakage was 0.49%. Leakage for pristine N95 FFRs is reported as
mean values and their standard errors ( $N = 3$ ).
\*The expectation values, confidence intervals and coefficients of determination are from power
regression performed on penetration measurements.

## Author contributions

P.Z.C. designed the study, performed experiments, analyzed results and prepared visualizations. A.N. and N.M. performed experiments. J.T.M., G.H.B., O.D.R. and F.X.G. supervised the research. P.Z.C. and F.X.G. wrote the manuscript with review from all authors.

## Acknowledgements

We would like to thank Jeffrey Sun, Febby Wong, Yang Ting Shek and Ayoob Ghalami at the University of Toronto for assistance with fit testing and procurement; Ronald Hofmann and Chengjin Wang at the University of Toronto for use of their fiber optic spectrometer and discussions; and Stephenie Naugler and William Lau at St. Michael’s Hospital for assistance with reprocessing instrumentation. This research was supported by the Natural Sciences and Engineering Research Council of Canada (NSERC), the University of Toronto COVID-19 Action Grant, the Hospital for Sick Children and Unity Health Toronto. P.Z.C. was supported by the NSERC Vanier Scholarship. F.X.G. was partially supported by the NSERC Senior Industrial Research Chair program.

## Notes

### Competing Interest Statement

The authors have declared no competing interest.

